# Sphenopalatine Ganglion Block for Post-Dural Puncture Headache: A Randomised Controlled Trial

**DOI:** 10.64898/2026.05.06.26352338

**Authors:** Gustavo Everardo-Salazar, Phabel A. López-Delgado, Mirna M. Delgado-Carlo

## Abstract

**Background:** Post-dural puncture headache (PDPH) affects up to 11.2% of patients after neuraxial anesthesia. The sphenopalatine ganglion block (SPGB) is a promising minimally invasive intervention, but high-quality randomised trial data are limited. We conducted a randomised controlled trial to evaluate the efficacy of SPGB compared to conservative management for PDPH.

**Methods:** Twenty-six patients with PDPH following accidental dural puncture with 17G Tuohy needles were randomised to conservative management (bed rest, hydration) or SPGB (bilateral intranasal 2% lidocaine). The sample size was calculated using a two-sided test for comparing two proportions, with *α* = 0.05 and 80% power, assuming a 60% difference in pain resolution between groups (80% vs 20%), yielding 12 patients per group; accounting for 10% attrition, 26 patients were enrolled. Pain intensity was measured using the Numeric Rating Scale (NRS, 0-10) at 30 minutes, 12 hours, and 24 hours. Secondary outcomes included rescue analgesia requirements, mobilization time, and adverse events.

**Results:** Recruitment, retention, and protocol adherence were 100%. At 30 minutes, all SPGB patients reported complete pain resolution (NRS=0) versus median NRS 3 (IQR 2) in controls (p<0.001). No SPGB patients required rescue analgesia or experienced adverse events. Conservative group patients had prolonged hospitalization (46%). Based on these findings, a sample size calculation for a future sham-controlled trial (90% power, *α* = 0.05) yields 120 participants (60/group).

**Conclusions:** SPGB provided rapid and complete pain resolution in all treated patients, with no adverse events and significantly earlier mobilization compared to conservative management. These findings support the conduct of a larger sham-controlled trial to confirm the efficacy of SPGB for PDPH.

**Trial registration:** ClinicalTrials.gov - NCT07494383 (retrospectively registered).

**Highlights:**

1. RCT demonstrates that SPGB provides complete pain resolution in all 13 treated patients at 30 minutes.
2. 100% recruitment, retention, and protocol adherence achieved (n=26).
3. All SPGB patients reported complete pain resolution (NRS=0) at 30 minutes.
4. No adverse events with SPGB; 46% of controls required prolonged hospitalization.
5. A future sham-controlled trial would require 120 participants (60/group) for 90% power.

## 1. Figure 1: Graphical Abstract – CONSORT

**Figure 1:**
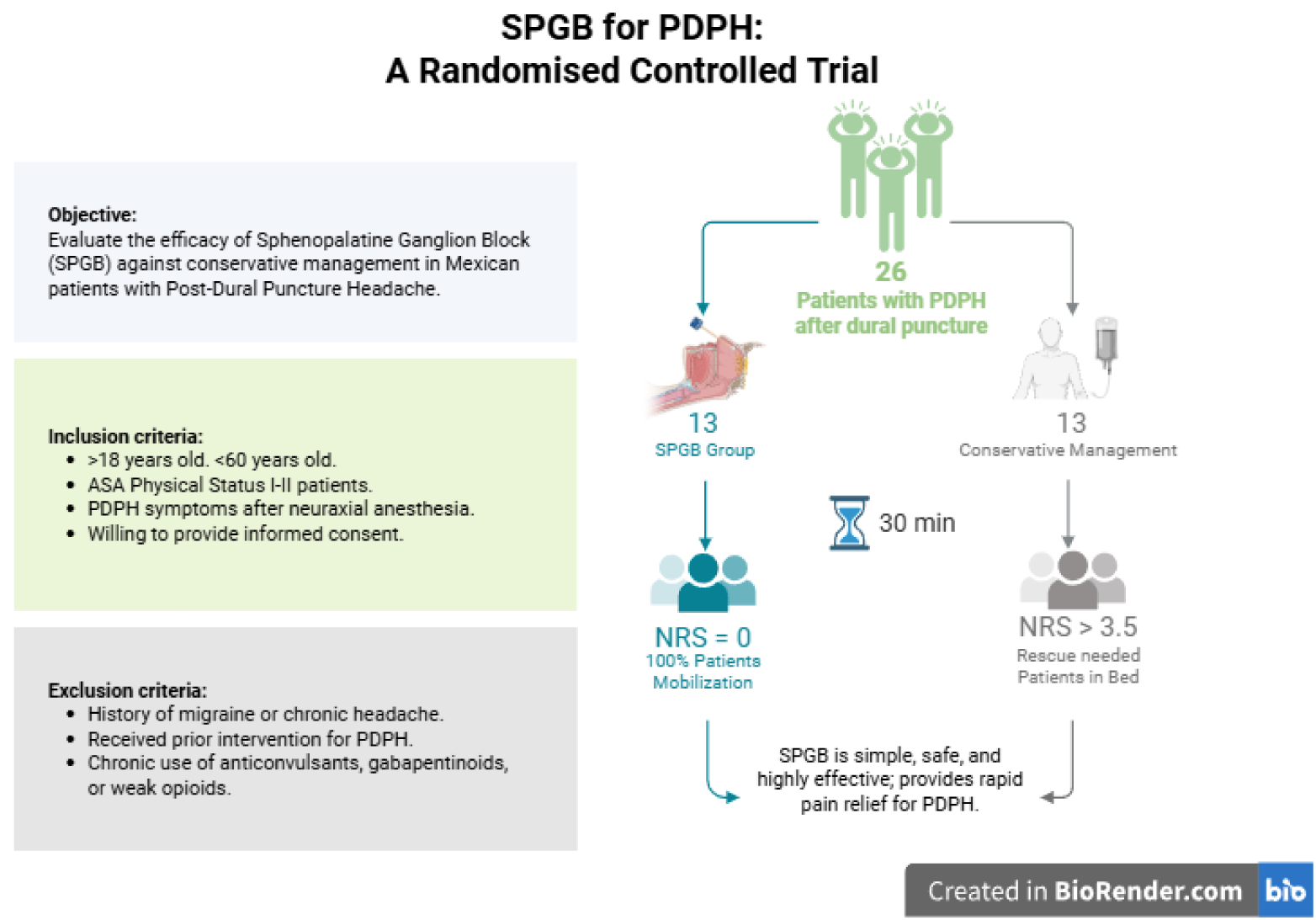
Graphical Abstract - CONSORT Comparison of sphenopalatine ganglion block (SPGB) versus conservative management for post-dural puncture headache (PDPH). (A) Randomization of patients with PDPH. (B) SPGB group shows rapid pain resolution (NRS=0 at 30 min) and immediate mobilization. (C) Conservative management group shows persistent pain, prolonged bed rest, and need for rescue analgesia.

## 2. Introduction

Post-dural puncture headache (PDPH) is a significant quality indicator in anesthesiology practice [1, 2]. In Mexico, various official standards (Normas Oficiales Mexicanas, NOM) have been established to homogenize, systematize, and update healthcare procedures, part of the “Health Sector Reform Programme” aimed at improving the quality of care. While these standards establish a comprehensive legal framework for anesthesiology practice, they do not specifically address the evaluation of quality indicators such as PDPH rates [3].

PDPH is the most common complication of neuraxial anesthesia, first described by August Bier in 1899 [4]. Its incidence varies between 1.5% and 11.2% following spinal anesthesia and 0.19% to 3.6% after epidural catheter insertion [5]. The most accepted pathophysiological theory suggests dural puncture causes decreased cerebrospinal fluid (CSF) pressure, triggering reflexive meningeal vasodilation [6, 7].

The primary goal of treatment is prevention, but when PDPH occurs, current management ranges from conservative measures to invasive interventions. Although the epidural blood patch (EBP) remains the gold standard for refractory cases, its use is limited by its invasive nature, requirement for specialist expertise, and non-negligible risk of complications, including repeated dural puncture, infection, and back pain. These factors can lead to delayed treatment, particularly in resource-constrained settings. A simple, minimally invasive, and rapid alternative that could be applied earlier in the treatment algorithm is therefore needed [8, 9]. Most authors therefore suggest waiting 24–48 hours before implementation, as approximately 85% of cases resolve spontaneously within this period [2, 10, 11]. Conventional conservative measures during this interval include bed rest, prone positioning, and aggressive hydration, though evidence supporting these practices is limited [10, 12].

The sphenopalatine ganglion block (SPGB) has recently emerged as a promising intervention for PDPH. Its mechanism involves blockade of parasympathetic outflow to cerebral vasculature, inhibiting release of pro-inflammatory neuropeptides [7, 10, 11, 13–18].

Given the limitations of conventional treatment and the emerging evidence for SPGB, we conducted a randomised controlled trial to evaluate the efficacy of SPGB compared to conservative management for PDPH following accidental dural puncture with large-bore (17G) Tuohy needles. The sample size was calculated using a two-sided test for comparing two proportions, with *α* = 0.05 and 80% power, assuming a 40% difference in pain resolution between groups (80% in the SPGB group vs 40% in the conservative group). This yielded 22 patients per group; accounting for 15% attrition, a total of 26 patients were enrolled. We hypothesized that SPGB would provide superior pain relief and earlier mobilization compared to conservative management.

## 3. Methods (Fig. 1)

### 3.1. Study Design and Setting

This was a single-centre, prospective, randomised, controlled, parallel-group trial conducted at the Hospital Regional “General Ignacio Zaragoza”, ISSSTE. The study was approved by the Institutional Bioethics and Research Committee (Reference Number: A2024-1014) and was conducted in accordance with Good Clinical Practice standards and Mexican health research regulations.

### 3.2. Sample Size Calculation

The sample size was calculated using a two-sided test for comparing two proportions, with *α* = 0.05 and 80% power. Assuming a 60% difference in pain resolution between the SPGB group (80%) and the conservative management group (20%), the required sample size was 12 patients per group. Accounting for a 10% attrition rate, the target sample size was set at 26 patients (13 per group).

### 3.3. Participants

We enrolled patients aged 18–55 years with ASA physical status I–II who developed PDPH symptoms following documented or suspected dural puncture during neuraxial anesthesia. Patients were enrolled within 24 hours of dural puncture. PDPH was diagnosed clinically as headache occurring within 5 days of neuraxial puncture with postural characteristics (worsening upright, relief supine), consistent with ICHD criteria. Cases could include associated neck stiffness and auditory symptoms. All cases in this study resulted from accidental dural puncture during epidural procedures performed with 17G Tuohy needles. This represents a high-risk population for severe PDPH given the large needle diameter. Written informed consent was obtained from all participants prior to enrolment.

**Exclusion Criteria:** History of migraine or chronic headache; received prior intervention for PDPH; chronic use of anticonvulsants, gabapentinoids, or weak opioids.

**Elimination Criteria:** Patient withdrawal of consent; development of side effects requiring further interventional management.

### 3.4. Randomization and Interventions

Participants were randomly allocated using a computer-generated random number table. To minimize the time spent in pain before receiving treatment, interventions were initiated immediately after randomization without formal baseline pain assessment.

**Control Group (Conservative Management):** Patients were immediately positioned supine then prone, with bathroom privileges at bedside, and received aggressive intravenous hydration: IV 0.9% saline at 125 mL/hr plus a minimum of 3 L oral fluids per 24 hours. This conservative management protocol reflected standard institutional practice at ISSSTE and Mexican public hospitals, in accordance with applicable national health regulations. Pharmacological adjuncts such as caffeine and gabapentin were intentionally excluded to isolate the specific effect of SPGB and avoid confounding.

**Intervention Group (SPGB):** With the patient supine with slight cervical extension, cotton-tipped applicators saturated with 2 mL of 2% lidocaine were inserted bilaterally at a 45° angle to the hard palate, advanced along the superior border of the middle turbinate to the posterior nasopharyngeal wall, and left in place for 15–20 minutes before removal.

**Time Zero Definition:** The first pain assessment (T0) was conducted immediately following completion of the intervention procedure in both groups. For the SPGB group, this occurred upon removal of the cotton applicators (approximately 20 minutes post-randomisation). For the conservative group, this occurred shortly after positioning and initiation of hydration (approximately 5 minutes post-randomisation).

### 3.5. Outcomes and Data Collection

This was a single-blind trial. Patients and clinicians could not be blinded due to the nature of the interventions. The outcome assessor collecting NRS scores was instructed to remain blinded to group allocation and was not involved in treatment delivery.

Pain intensity was measured using the Numeric Rating Scale (NRS, 0–10) immediately post-intervention (T0), at 30 minutes (T1), 12 hours (T2), and 24 hours (T3) post-intervention. Other measures included time to mobilisation from bed, need for rescue analgesia (tramadol), and adverse events.

### 3.6. Statistical Analysis

Data were analysed using R statistical software (version 4.3.1). Between-group differences in NRS scores at each timepoint were analysed using the Mann–Whitney U test. Effect sizes are expressed as rank-biserial correlation coefficients (*r*) with 95% confidence intervals, calculated using the *ggstatsplot* package [19]. A two-sided *p <* 0.05 was considered statistically significant. No imputation was performed for missing data.

### 3.7. Ethical Considerations

The study adhered to biomedical ethics principles: autonomy (written informed consent), beneficence (intervention aimed to provide faster symptomatic relief), non-maleficence (strict aseptic technique and single-use materials), and justice (equitable participant selection). Epidural blood patch was available as rescue therapy after 24 hours, but none was required.

### 3.8. Safety Considerations

Lidocaine use was approved by the FDA and COFEPRIS with a Category B safety classification. Comprehensive patient assessment preceded the procedure, and drug preparation/administration followed strict aseptic technique with single-use materials.

### 3.9. Trial Registration

This trial was prospectively registered with the Instituto de Seguridad y Servicios Sociales de los Trabajadores del Estado (ISSSTE) under registration number **ISSSTE RPI #408-2024**. The trial was retrospectively registered on ClinicalTrials.gov (ID: **NCT07494383**). All outcomes are reported in accordance with CONSORT guidelines.

## 4. Results

### 4.1. Recruitment and Retention

A total of 26 patients who developed PDPH following neuraxial anesthesia were enrolled and randomised. All 26 randomised participants (13 per group) received their allocated intervention and completed follow-up, yielding a retention rate of 100%. Protocol adherence was 100%. No participants were lost to follow-up or requested withdrawal. Data completeness for the primary clinical measure (NRS) was 100% at all timepoints.

### 4.2. Patient Characteristics

Baseline characteristics are presented in Table **??**. The groups were comparable in age and ASA status, with a female predominance in both cohorts.

**Table 1:**
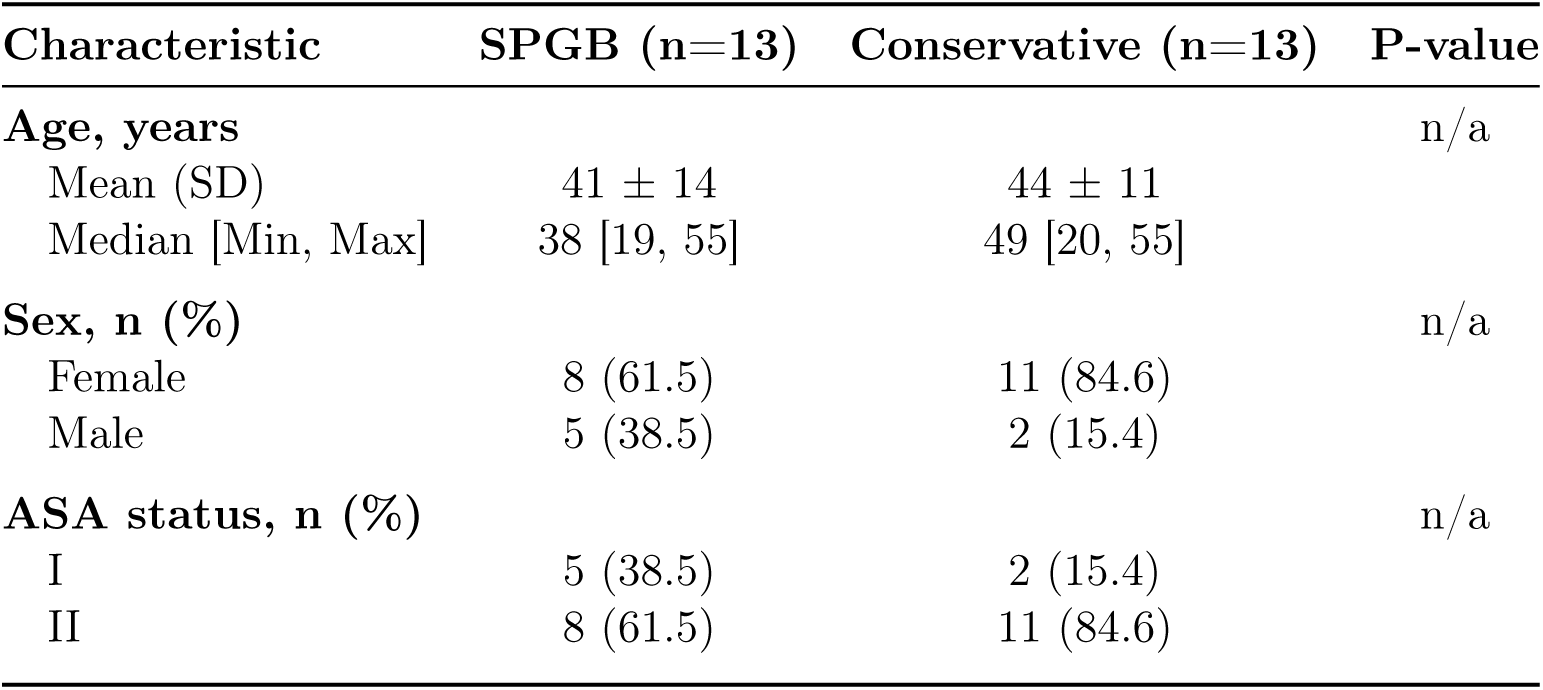
Demographic characteristics of study participants.

**Table 2:**
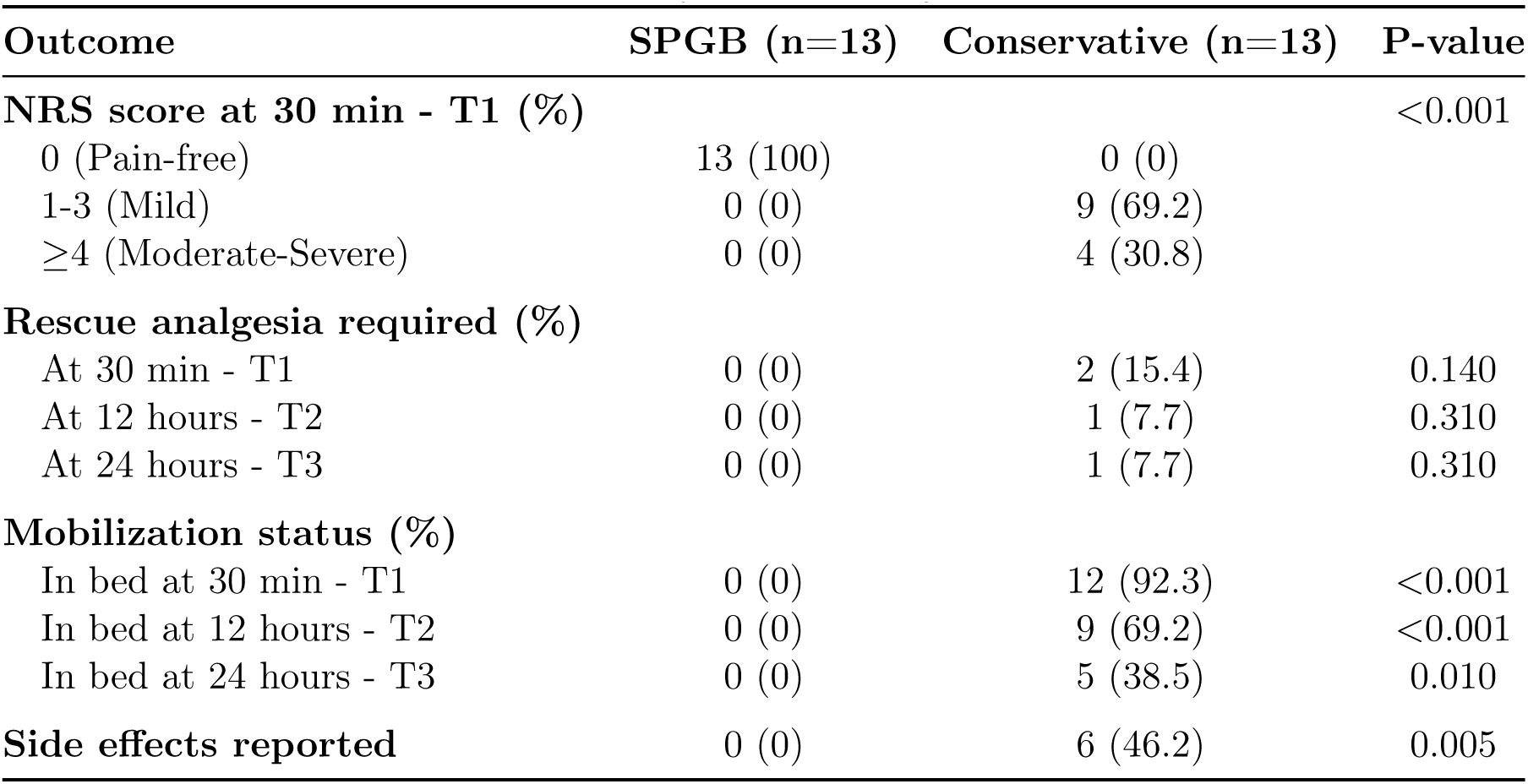
Clinical outcomes of study participants by Treatment Group.

### 4.3. Clinical Outcomes

#### 4.3.1. Pain Intensity

NRS scores over time are shown in Figure 2A and Table **??**. At T0 (immediately post-intervention), the SPGB group presented higher initial pain scores (mean NRS 4.8, median 4, IQR 2) compared to the conservative group (mean NRS 1.9, median 2, IQR 3). The interpretation of this timing difference is discussed in Section 5.3.

**Figure 2:**
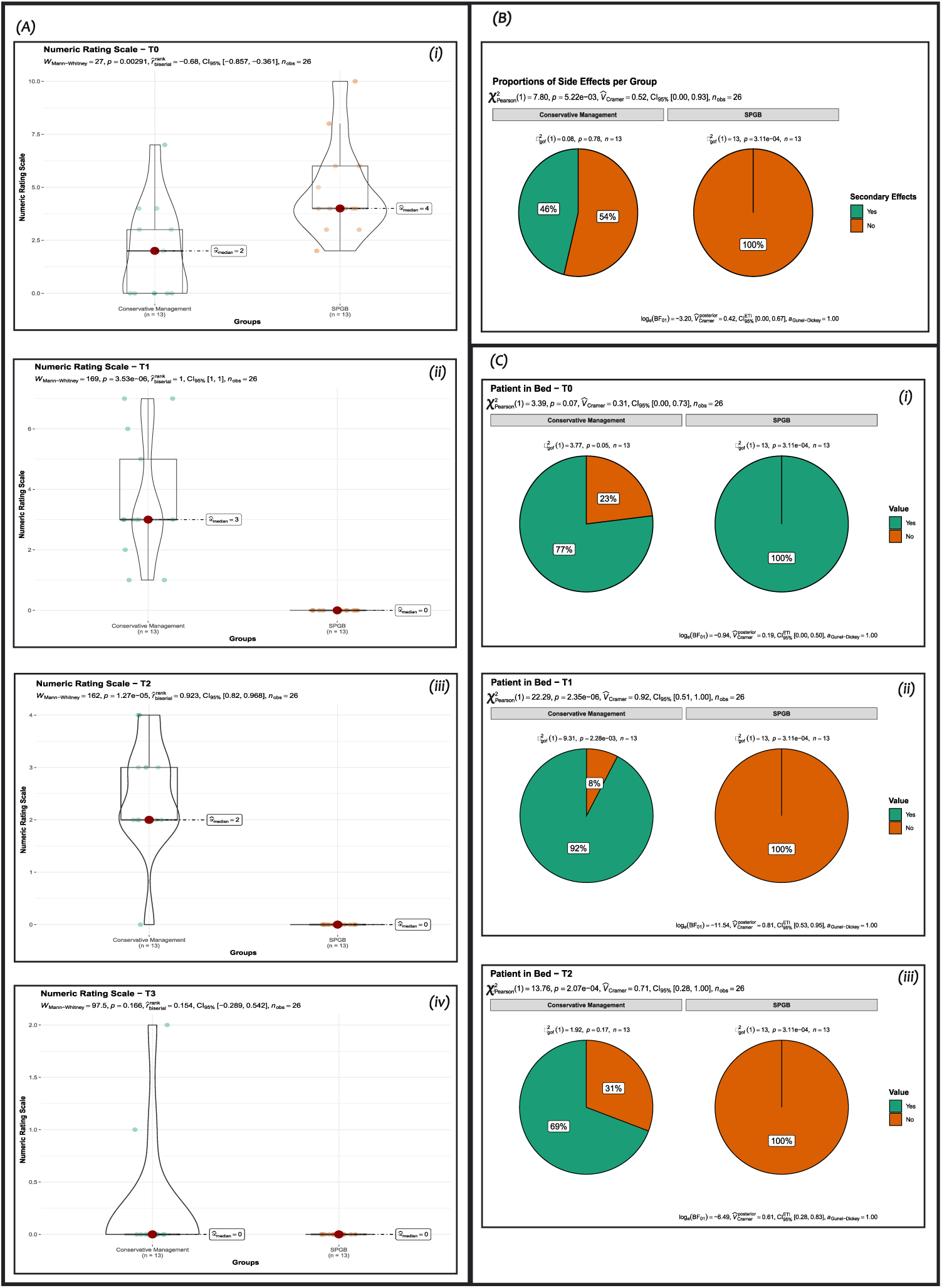
**A**) NRS scores over time for SPGB and conservative groups. Box plots show median, interquartile range, and range. **B)** Incidence of protocol-related side effects. **C)** Patient mobilisation status at each timepoint, showing proportion of patients remaining in bed.

At 30 minutes post-intervention (T1), the SPGB group showed complete pain resolution (mean NRS 0, median 0, IQR 0), while the conservative management group showed minimal improvement (mean NRS 3.6, median 3, IQR 2). The Mann–Whitney U test indicated a statistically significant difference (p < 0.001), with complete separation between groups resulting in a ceiling effect size of *r* = 1.0 (95% CI [1.0, 1.0]).

At later timepoints (T2, T3), differences between groups attenuated but remained apparent. The effect sizes at T2 (12 hours) and T3 (24 hours) were *r* = 0.93 (95% CI [0.83, 0.96]) and *r* = 0.15 (95% CI [-0.28, 0.54]), respectively (Table **??**).

#### 4.3.2. Adverse Events

A prolonged hospital stay was documented in 6 patients (46%) in the conservative group. No patient in the SPGB group experienced anosmia or any other reported side effect (p = 0.005) (Figure 2B). No serious adverse events were observed in either group.

#### 4.3.3. Rescue Analgesia and Mobilisation

No patient in the SPGB group required rescue analgesia (tramadol) at any time point. In the conservative management group, rescue analgesia was required in 2 patients (15.4%) at 30 minutes, decreasing over time, with one patient (7.7%) still requiring it at 24 hours (T3). Patients in the SPGB group were able to ambulate freely without PDPH symptoms at 30 minutes, whereas those receiving conservative management remained bedridden for significantly longer (Figure 2C, p < 0.001 at T1).

### 4.4. Post-hoc Sample Size Estimation

Based on these findings, a sample size calculation for a future multi-centre trial was performed. The primary outcome was specified as the NRS score at 30 minutes post-intervention (T1). Using a clinically anchored approach with the Minimally Clinically Important Difference (MCID) for acute pain on an NRS (1.3 points), and the pooled standard deviation of NRS scores at T1 (2.31), the target effect size was Cohen’s *d* = 0.56. Using a two-sided *α* = 0.05 and 90% power for an independent samples t-test, the required sample size is 108 participants. Accounting for 10% attrition, the final recruitment target for a definitive multi-centre RCT is 120 participants (60 per group).

## 5. Discussion

This randomised controlled trial evaluated the efficacy of sphenopalatine ganglion block (SPGB) compared to conservative management for post-dural puncture headache (PDPH) following accidental dural puncture with large-bore (17G) Tuohy needles. The study demonstrated that SPGB provided rapid and complete pain resolution in all treated patients at 30 minutes, with no adverse events and significantly earlier mobilisation compared to conservative management.

### 5.1. Summary of Key Findings

All 13 patients (100%) in the SPGB group achieved complete pain resolution (NRS=0) at 30 minutes post-intervention, compared to a median NRS of 3 (IQR 2) in the conservative group (p < 0.001). No patient in the SPGB group required rescue analgesia or experienced adverse events, while 46% of conservative group patients had prolonged hospitalisation. Patients in the SPGB group were able to ambulate freely at 30 minutes, whereas conservative management patients remained bedridden for significantly longer.

### 5.2. Comparison with Previous Studies

The dramatic early response to SPGB is consistent with the known pharmacodynamic profile of lidocaine and the proposed mechanism of SPGB, which involves blockade of parasympathetic outflow to cerebral vasculature and inhibition of pro-inflammatory neuropeptides [7, 10, 11]. In contrast, conservative management showed minimal early improvement, aligning with previous reports on the limitations of bed rest and hydration [9, 10].

An important feature of our study population is that all cases resulted from accidental dural puncture with large-bore (17G) Tuohy needles during epidural procedures. This represents a particularly challenging clinical scenario, as PDPH incidence and severity are substantially higher following large-needle punctures compared to routine spinal anesthesia with smaller gauges. The fact that SPGB achieved complete pain resolution in this high-risk population suggests it may be even more effective for routine PDPH from smaller spinal needles.

Our findings differ from the high-quality blinded RCT by Jespersen et al. [12], which compared SPGB to sham and found no difference in headache severity at 24 hours. Several methodological differences may explain this discrepancy. First, their primary endpoint assessed lasting effects at a timepoint (24 hours) when spontaneous resolution may obscure treatment differences; our acute assessment (30 minutes) was designed to capture the pharmacodynamic window of SPGB. Second, their population included mixed needle gauges (predominantly smaller spinal needles), whereas our uniform 17G cohort represents more severe PDPH likely to benefit from intervention. Critically, our open-label design introduces performance and detection bias not present in their blinded trial, meaning placebo effects cannot be excluded as contributing to our dramatic findings. A meta-analysis by Hung et al. [18] similarly concluded negatively regarding SPGB efficacy for PDPH; however, as with Jespersen et al., the pooled studies assessed later endpoints and mixed needle-gauge populations, limiting direct comparability with our acute, uniform 17G cohort.

### 5.3. Interpretation of T0 Measurements

The higher pain scores observed in the SPGB group at T0 require careful interpretation. As no pre-randomisation assessment was performed, T0 represents the first post-intervention measurement rather than a true baseline — the conservative group was assessed approximately 5 minutes after positioning and hydration initiation, while the SPGB group was assessed immediately after 15–20 minutes of applicator contact. The apparent difference at T0 may therefore reflect early pharmacodynamic activity of lidocaine during applicator placement, differential measurement timing, or natural pain fluctuation, rather than a pre-existing between-group difference at enrolment.

### 5.4. Strengths and Limitations

This study has several strengths. The use of a uniform, high-risk cohort (all patients with 17G Tuohy needle punctures) provides a well-defined population with clinically significant PDPH. The sample size was calculated a priori to detect a clinically meaningful difference in pain resolution. Protocol adherence and retention were 100%, supporting the feasibility of this intervention in clinical practice.

Several limitations must be acknowledged. The single-centre design and small sample size limit generalisability and precision of effect estimates. The absence of a pre-randomisation pain assessment was an ethical choice to minimise treatment delay, but means we cannot formally verify group equivalence at enrolment. The intervention could not be blinded to patients or clinicians, introducing potential performance bias; however, a blinded outcome assessor was used for pain scoring. The conservative management protocol, while based on traditional practice, may have been overly restrictive (strict bed rest with minimal mobilisation) [10, 12], potentially widening the apparent treatment effect. Finally, the retrospective registration of the trial is a limitation, though this does not affect the integrity of the data.

### 5.5. Implications for Future Research

The dramatic early response to SPGB in this high-risk cohort supports the conduct of a larger, sham-controlled, multi-centre RCT to confirm efficacy. Based on the observed effect size, a definitive trial would require 120 participants (60 per group) for 90% power. Future trials should incorporate pre-randomisation baseline assessments, sham-controlled blinding, and pragmatic conservative management to strengthen internal validity and generalisability. Stratification by needle gauge and puncture type would further clarify the patient populations most likely to benefit from SPGB.

### 5.6. Conclusions

In this randomised controlled trial, SPGB provided rapid and complete pain resolution in all treated patients with PDPH following accidental dural puncture with large-bore needles, with no adverse events and significantly earlier mobilisation compared to conservative management. These findings support the conduct of a larger sham-controlled trial to confirm the efficacy of SPGB for PDPH.

## 6. Authors’ Contributions

GE: conceptualization, data curation, formal analysis, funding acquisition, investigation, methodology, project administration, resources, supervision, validation, writing- reviewing and editing; PL: data curation, formal analysis, methodology, project administration, software, supervision, validation, visualization, writing- reviewing and editing; MD: conceptualization, funding acquisition, investigation, methodology, project administration, resources, supervision, validation, writing- reviewing and editing.

All authors have read and approved the final manuscript.

## Data Availability

All data produced in the present study are available upon reasonable request to the authors. Deidentified individual participant data will be made available following article publication, on GitHub <https://github.com/phabel-LD>, along with all analysis code.

## Acknowledgments

The authors acknowledge the Instituto de Seguridad y Servicios Sociales de los Trabajadores del Estado Hospital Regional “General Ignacio Zaragoza”, its Department of Anesthesiology, Dr. Miguel Pineda Sánchez, M.D.; and the Universidad Nacional Autónoma de México.

## 9. Data Availability

Deidentified individual participant data will be made available following article publication, on GitHub <https://github.com/phabel-LD>, along with all analysis code.

